# Post lockdown COVID-19 seroprevalence and circulation at the time of delivery, France

**DOI:** 10.1101/2020.07.14.20153304

**Authors:** Jérémie Mattern, Christelle Vauloup-Fellous, Hoda Zakaria, Alexandra Benachi, Julie Carrara, Alexandra Letourneau, Nadège Bourgeois-Nicolaos, Daniele De Luca, Florence Doucet-Populaire, Alexandre J. Vivanti

**Affiliations:** Division of Obstetrics and Gynecology, Antoine Béclère Hospital, Paris Saclay University, AP-HP, Clamart, France; Division of Virology, Paul Brousse Hospital, Paris Saclay University, AP-HP, INSERM U1193 Villejuif, France; Division of Microbiology, Antoine Béclère Hospital, Paris Saclay University, AP-HP, Clamart, France; Division of Pediatrics and Neonatal Critical Care, Antoine Béclère Hospital, Paris Saclay University, AP-HP, Clamart, France

## Abstract

**Background:** To fight the COVID-19 pandemic, lockdown has been decreed in many countries worldwide. The impact of pregnancy as a severity risk factor is still debated, but strict lockdown measures have been recommended for pregnant women.

**Objectives:** To evaluate the impact of the COVID-19 pandemic and lockdown on the seroprevalence and circulation of SARS-CoV-2 in a maternity ward in an area that has been significantly affected by the virus.

**Study design:** Prospective study at the Antoine Béclère Hospital maternity ward (Paris area, France) from May 4 (one week before the end of lockdown) to May 31, 2020 (three weeks after the end of lockdown). All patients admitted to the delivery room during this period were offered a SARS-CoV-2 serology test as well concomitant SARS-CoV-2 RT-PCR on a nasopharyngeal sample.

**Results:** A total of 249 women were included. Seroprevalence of SARS-CoV-2 was 8%. The RT-PCR positive rate was 0.5%. 47.4% of the SARS-CoV-2-IgG-positive pregnant women never experienced any symptoms. A history of symptoms during the epidemic, such as fever, myalgia and anosmia, was suggestive of previous infection.

**Conclusions:** Three weeks after the end of lockdown, SARS-CoV-2 infections were scarce in our region. A high proportion of SARS-CoV-2-IgG-negative pregnant women must be taken into consideration in the event of a resurgence of the pandemic in order to adapt public health measures to reduce exposure to the virus, such as social distancing and teleworking for this specific population.

## Introduction

The coronavirus disease 2019 (COVID-19) pandemic hit France at the end of February 2020. To date (June 15, 2020), more than 7,000,000 cases and 400,000 deaths have occurred worldwide.[1] France, and more particularly the Paris area, was one of the regions of the world most affected by the pandemic. In order to slow down progression of the pandemic, lockdown has been decreed in many countries worldwide. French lockdown started on March 17, 2020, and ended on May 11, 2020.

Pregnancy as a severity risk factor of severe acute respiratory syndrome coronavirus 2 (SARS-CoV-2) infection is still debated, even though the third trimester seems to be a consensual risk factor[2–7]. The main goal of this study was to evaluate the impact of the first wave of the COVID-19 pandemic in our region and the effect of lockdown on seroprevalence in our maternity ward, which has been significantly affected by the virus (more than 40 confirmed infections among pregnant women between March 12 and April 20).

## Materials and Methods

We conducted a prospective study at the Antoine Béclère Hospital maternity ward (Paris area, France) from May 4 (one week before the end of lockdown) to May 31, 2020 (three weeks after the end of lockdown). All patients admitted to the delivery room during this period were offered a SARS-CoV-2 serology test as well as SARS-CoV-2 RT-PCR on a nasopharyngeal sample. A questionnaire was distributed to all included women, who were asked about symptoms (fever, cough, dyspnea, myalgia, anosmia, diarrhea and rash) affecting them and/or close relatives (household residents) from January 1 to their delivery, and if there were symptoms they were asked for the date of onset. Patients were also asked whether they strictly adhered to the rules of lockdown and social distancing.

### Serology testing

Serum samples were run on the Abbott Architect instrument using the Abbott SARS- CoV-2 IgG assay (Abbott, Sligo, Ireland) following the manufacturer’s instructions. The assay is a chemiluminescent microparticle immunoassay for detection in human serum or plasma of IgG against the SARS-CoV-2 nucleocapsid protein. Based on our local evaluation of 500 serum samples (data not published), the result was considered negative if the IgG index value was < 0.8, equivocal if the IgG index was between 0.8 and 1.39, and positive if the IgG index value was ≥ 1.40 (0.40 is the Abbott determined positivity cut-off).

### Molecular testing

Nasopharyngeal swabs were collected from all the patients. Samples were tested by using the GeneFinder^™^ COVID-19 Plus Real*Amp* Kit on the ELITe InGenius⟶ Platform, or the Xpert Xpress SARS-CoV-2 assay (Cepheid), or the GenMark ePlex SARS-CoV-2 assay, depending on the time of day.

The GeneFinder assay is a molecular in vitro test utilizing one-step reverse transcription real-time PCR to detect the RNA-dependent RNA polymerase, the envelope gene (E) and the nucleocapsid gene (N), as described in international guidelines (Corman *et al*.[8])

The Xpert Xpress SARS-CoV-2 assay (Cepheid) and the GenMark ePlex SARS-CoV- 2 assay are sample-to-answer molecular diagnostic platforms. The former is based on the detection of 2 gene targets, N2 and E, according to an algorithm in parallel with the sample process control, and the latter is based on the detection of the N gene of SARS-CoV-2.

All data (clinical, laboratory, from both mothers and newborns) were prospectively collected from medical records.

All statistical analyses were performed using R software (Version 3.5.1, R Core Team [2018] https://www.R-project.org/). Continuous variables are expressed as medians with interquartile range (IQR). Categorical variables are expressed as numbers with percentages. A two-tailed Mann-Whitney U test was used for statistical analysis of continuous variables. Fisher’s exact test was used for statistical analysis of categorical variables. Statistical significance was considered with a p<0.05.

This study was approved by the institutional review board of the French College of Obstetricians and Gynecologists (2020-OBST-0408) and was performed in accordance with the Declaration of Helsinki. All patients gave written consent. All data were de-identified to ensure patient privacy and confidentiality.

## Results

During the study period, SARS-CoV-2 serology testing was offered to all 272 patients admitted to the delivery room (figure 1). A total of 249 (91.5%) serology results were available (22 women did not give consent and one missing result). Seroprevalence was 8% (n=20/249). The main characteristics of the women and of their pregnancies and newborns are shown in Table 1. All patients were asymptomatic on admission to the delivery room. There were no significant differences between the groups of SARS-CoV-2-IgG-positive and SARS-CoV-2-IgG-negative women. Among women admitted for delivery, 190 had a SARS-CoV-2 RT-PCR test during the study period (59 women declined testing and one missing result). Exhaustivity was therefore of 69.9% (190/272). Only one (0.5%) test was positive and this patient remained asymptomatic during delivery and early post-partum. This positive test was performed on May 25, 2020, using the Xpert Xpress assay, with detection of the N2 gene at a CT value of 41.4, which corresponds to a very low viral load, which may be related to an old infection. The serology was positive. The patient and her spouse reported symptoms such as headache, asthenia and anosmia as of March 10, 2020. Her newborn tested negative (negative SARS-CoV-2 RT-PCR performed on nasopharyngeal swabs at birth and at three days of life).

**Table 1:** Maternal, obstetric and neonatal characteristics according to SARS-CoV-2 serological status for women admitted for delivery.

|  |  |  |  |
| --- | --- | --- | --- |
|  | **IgG-negative**<br>**n=229** | **IgG-positive**<br>**n=20** | **p** |
| Age, years, median [IQR] | 33 [29-36] | 31 [30.5-37] | 0.88 |
| Nullipara, n (%) | 106 (46.3) | 12 (60) | 0.25 |
| BMI, kg/m<sup>2</sup>, median [IQR] | 23 [21-27] | 25,5 [23-28.2] | 0.07 |
| History of preexisting: |  |  |  |
| - Diabetes mellitus, n (%) | 2 (0.9) | 0 (0) | 1 |
| - Chronic hypertension, n (%) | 6 (2.6) | 0 (0) | 1 |
| - Tobacco use, n (%) | 44 (19.2) | 2 (10.6) | 0.54 |
| - Asthma, n (%) | 13 (5.7) | 0 (0) | 0.6 |
| Gestational age at delivery, week median [IQR] | 39.6 [38.2-40.7] | 39.7 [38.2-40.3] | 0.50 |
| Delivery before 37 WG, n (%) | 26 (11.4) | 4 (21.1) | 0.26 |
| Birthweight, g median [IQR] | 3241 [2829-3585] | 3213 [2853-3471] | 0.62 |
| Infected neonates, n (%) | 0 (0) | 0 (0) |  |

**Figure 1:** Flow chart of the study – Universal serology screening for SARS-CoV-2 among a cohort of women admitted for delivery.

Analysis of the available questionnaires (n=220/249; 88.3%) showed that 47.4% (9/19) of the SARS-CoV-2-IgG-positive women reported being asymptomatic throughout the first wave of the pandemic. SARS-CoV-2-IgG-positive women reported more symptoms compared to SARS-CoV-2-IgG-negative women (Table 2), such as fever (n=3 (15.8%) versus n=3 (1.5%); OR=12.9 (95% CI 1.49-97.52); p=0.009), myalgia (n=7 (36.8%) versus n=14 (7%); OR=8.39 (95% CI 2.38-25.46); p<0.001) and anosmia (n=6 (31.6%) versus n=3 (1.5%); OR=29.2 (95% CI 5.52-201); p<0.001). There was no difference regarding the symptoms of cough, dyspnea, diarrhea and rash. All women except one reported they had strictly respected lockdown rules.

**Table 2:** Reported signs and symptoms according to SARS-CoV-2 serological status for women admitted for delivery.

|  |  |  |  |  |
| --- | --- | --- | --- | --- |
|  | IgG-negative<br>n=201 | IgG-positive<br>n=19 | OR (95% CI) | p |
| Fever, n (%) | 3 (1.5) | 3 (15.8) | OR=12.9 (1.49-97.52) | 0.009 |
| Cough, n (%) | 13 (6.5) | 2 (10.5) | OR=1.69 (0.17-8.52) | 0.62 |
| Dyspnea, n (%) | 12 (6) | 1 (5.3) | OR=0.87 (0.02-6.6) | 1 |
| Myalgia, n (%) | 14 (7) | 7 (36.8) | OR=8.39 (2.19-25.46) | <0.001 |
| Anosmia, n (%) | 3 (1.5) | 6 (31.6) | OR=29,2 (5.52-201) | <0.001 |
| Diarrhea, n (%) | 21 (10.4) | 1 (5.3) | OR=0.47 (0.01-3.34) | 0,7 |
| Rash, n (%) | 5 (2.5) | 1 (5.3) | OR=2.16 (0.04-20.9) | 0.42 |

## Discussion

SARS-CoV-2 screening (serology and RT-PCR on nasopharyngeal secretions) of all women at delivery showed that: 1- the seroprevalence of SARS-CoV-2 infection was 8% and most of the pregnant women remained uninfected; 2- half of the SARS-CoV-2-IgG-positive pregnant women were completely asymptomatic; 3- the virus was no longer circulating up to three weeks after the end of the lockdown.

In our maternity ward, implementation and generalized respect of lockdown rules and social distancing allowed for a drastic reduction in the number of confirmed cases. Pregnant women reported full lockdown compliance from March 17, 2020 until delivery. Despite the end of lockdown and the resumption of activity for members of the family circle, the rate of positive results by RT-PCR remained very low. In addition, reported symptoms such as fever, myalgia and anosmia were linked with previous infection with the SARS-CoV-2 virus.

The seroprevalence of our cohort was similar to that recently observed in the general population[9,10]. To date, it has not been assessed whether SARS-CoV-IgG, detected with currently available ELISA assays, are neutralizing antibodies and consequently confer protection. However, it is of major importance to have an estimation of the rate of pregnant women who have not already been infected, in order to implement adapted measures in the event of a second outbreak, such as social distancing and teleworking at least for this specific population [11,12].

One could argue that the study period would need to be longer in order to fully estimate the effect of the end of the lockdown and the risk of a resumption of the pandemic (so-called second wave). However, we believe that our study provides an important snapshot at a given point in time of the exposure to COVID-19 in an at-risk population in a region highly exposed to the virus.

## Data Availability

The datasets generated during and/or analyzed during the current study are available from the corresponding author on reasonable request.

## Acknowledgments

We would like to thank David Marsh for language editing.

